# Depression, Anxiety and Stress among the Caregivers of Clubfoot Patients

**DOI:** 10.1101/2025.08.01.25332806

**Authors:** Mahema Akter, Zahid Bin Sultan Nahid, Shahid Afridi, Shahnaz Begam, Muhammad Shahidul Islam, Md Furatul Haque

**Author notes:** **Corresponding author** Shahid Afridi, Lecturer, Department of Physiotherapy, SAIC College of Medical Science and Technology (SCMST), Dhaka, Bangladesh. **IRB**-*SCMST/BPT/IRB-24/05-07/03.

## Abstract

**Background:** Congenital talipes equinovarus (CTEV), often known as clubfoot, is a common birth defect of the bones and muscles, especially in places with few resources like South Asia.

**Objective:** To determine the prevalence of stress, anxiety, and depression in parents of children receiving Ponseti treatment for congenital talipes equinovarus (CTEV), as well as the sociodemographic correlates of these symptoms.

**Materials and methods:** From September 2023 to August 2024, a descriptive cross-sectional study was done at the SAIC College of Medical Science and Technology in Dhaka. Eighty-five caregivers filled out a standard questionnaire that included the DASS-21 scale. They also wrote down information on comorbidities, clinical features, and sociodemographics. We used SPSS version 22.0 to do the statistical analysis. Chi-square tests looked at how independent factors were related to psychological distress. We used a significance level of p < 0.05.

**Results:** 50.6% of the 85 people who took part said they were anxious (48.8% said it was very bad), 44.7% said they were depressed (26.3% said it was very bad), and 28.2% said they were stressed (45.8% said it was very bad). There was a strong link between stress and the weight of a newborn (p=0.013), the way the baby was born (p=0.056), the age of the caregiver (p=0.053), and the caregiver’s education (p=0.029). Anxiety was only linked to monthly income (p=0.009). One of the main worries was money, specifically how much it costs for families in remote areas to commute. A lot of kids were in the bracing period and had bilateral clubfoot. Most of the caretakers were stay-at-home moms.

**Conclusion:** The mental health of the caregivers of CTEV patients is significantly impacted by the clinical and economical difficulties. Integrating psychosocial support and moving services away from central locations may help caregivers deal with stress. Future studies should look at cognitive and sleep issues using random sampling and larger sample sizes to improve treatment methods and make them more generalizable.

## Background

Congenital talipes equinovarus (CTEV), also known as clubfoot, is one of the most common musculoskeletal birth defects. About 1 to 2 out of every 1,000 live newborns around the world have it [1]. It happens more often in places with few resources, like South Asia (3.4 out of every 1,000 births in Nigeria), than in Western countries (1.2 out of every 1,000 births in Europe and North America [2]. The Ponseti method, which is the best way to treat the disease, puts a lot of stress on parents and caregivers since it requires long periods of casting, bracing, and following caregiver instructions [3,4]. Studies show that the emotional toll of the diagnosis, the difficulty of the therapy, and the financial problems that caregivers of children with clubfoot face make them more likely to be depressed, anxious, and stressed [5, 7]. For example, a study in Turkey looked at how anxious and hopeless parents were before and after treatment. The results showed that their levels of anxiety and despair dropped dramatically, which shows that therapy can help people feel better mentally [6]. Fifteen and a half percent of caregivers in Nigeria said they were having emotional problems, and twelve percent said they were experiencing clinically significant parenting stress, which was worse for younger patients and throughout specific treatment phases [2].

Taking care of someone with clubfoot can have a big effect on their social and emotional lives in many ways, such as how their family works, how society sees them, and how much it costs. In Bangladesh and South Asia, where poverty and restricted access to healthcare are common, caregivers have to deal with extra costs including travel and lost income when they take loved ones to the hospital [7,8]. A study done in Ethiopia found that families of children with clubfoot had a lot of trouble with their money and relationships because taking care of these kids took so much time [7,9]. On the other hand, in high-income nations like the UK and South Africa, social support and ways to deal with stress were less effective, but this varied depending on income level and education [6,9]. For instance, South African parents felt they were stronger because of the support they had from their communities; meanwhile, UK caregivers said they found it harder when they were alone [6]. The goal of this work is to fill up the gaps in region-specific psychosocial assessments, especially in areas that haven’t been researched as much, such South Asia and Africa, where cultural and economic issues have a big impact on caregiver mental health [2,7]. Most of the existing research focuses on clinical outcomes, which means it doesn’t look at the connection between treatment adherence and parents’ mental health [10, 11]. Brazilian and Indian research, for example, show that not following through with bracing is commonly associated to stress on the caregiver, lack of knowledge, and the child’s suffering [12,13]. People could get past these mental blocks and do better on their tests and follow the rules better if they had counseling and support, among other targeted interventions [3,14].

This paper wants to put together research from all across the world about caregiver mental health by focusing on incidence rates, geographical variances, and specific solutions. Using data from places like Bangladesh, South Asia, and Western countries will provide us a full picture of how to make healthcare policies that will make life easier for caregivers and improve the effectiveness of treatment.

## Method

This descriptive cross-sectional study was to find out how much stress, anxiety, and sadness the people who care for clubfoot sufferers are going through. It was housed in the SAIC College of Medical Science and Technology (SCMST) in Mirpur, Dhaka, from September 2023 until August 2024. The data came from the Trauma Center Bangladesh (Shyamoli, Mirpur Road), BIRDEM General Hospital 2 (Mother and Child), and CRP (Mirpur-14). The study group was made up of caretakers of children between the ages of 18 and 50 who were getting Ponseti treatment for congenital talipes equinovarus (CTEV). People who were sick or had trouble thinking were not included. With a 16.8% prevalence and a 5% margin of error, only 85 caregivers out of the 215 total sample size were able to be reached. A pre-tested, standardized questionnaire was used for the in-person interviews. The tool has four parts: sociodemographic data, information about clubfoot, information about co-morbidity, and the DASS-21 scale for measuring stress, anxiety, and depression. Using established cut-off scores, the results were put into groups of normal, mild, moderate, severe, and very severe. We graded each DASS-21 item on a scale of 1 to 4. There were six trips to the medical facilities on the list to gather data. Following the requirements of the BMRC and the WHO, all participants gave their informed consent after the Ethical Review Board at SCMST approved it. The study put a lot of emphasis on voluntary involvement and keeping things private. We coded, changed, and put the data into Microsoft Excel 2016 and SPSS version 22.0 with different settings so that we could analyze it correctly. Chi-square tests and descriptive statistics (means, percentages, and frequencies) helped people comprehend how variables are related. Putting the results in tables and graphs helped make things clearer. Participants could leave at any time, and no intrusive procedures were done.

## Results

The study investigated the answers to 85 questionnaires to find out how stressed, anxious, and depressed people who look for clubfoot (CTEV) patients are. Most of the caregivers were women (98.8%), homemakers (95.3%), and older than 23.

| Variables | Category | Frequency | Percentage (%) |
| --- | --- | --- | --- |
| Age of Caregiver | 18-22 years | 17 | 20.0 |
|  | 23 -25 years | 19 | 22.4 |
|  | 26 -30 years | 23 | 27.1 |
|  | > 30 year | 26 | 30.6 |
| Age of caregivers overall (Mean ±SD) = (2.68 ±1.115) | | | |
| Age of Children | 1-4 month | 19 | 22.4 |
|  | 5-11 month | 21 | 24.7 |
|  | 12-23 month | 15 | 17.6 |
|  | >23 month | 30 | 35.3 |
| Age of Children overall (Mean ±SD) = (2.66 ±1.181.) | | | |
| Educational status of caregiver | Non formal | 1 | 1.2 |
|  | Primary | 27 | 31.8 |
|  | Secondary | 29 | 34.1 |
|  | Bachelor of degree | 28 | 32.9 |
| Educational status of caregiver overall (Mean ±SD) = (1.99 ±0.838) | | | |
| Occupation of caregiver | Homemaker | 81 | 95.3 |
|  | Others | 4 | 4.7 |
| Occupation of caregiver overall (Mean ±SD) = (1.09 ±0.426) | | | |
| Monthly family income | 10000-15000 | 6 | 7.1 |
|  | 16000-30000 | 34 | 40 |
|  | 31000-40000 | 11 | 12.9 |
|  | >40000 | 34 | 40 |
| Monthly family income overall (Mean ±SD) = (2.86 ±1.04) | | | |

Forty percent of them made more than 40,000 BDT a month, and their education level ranged from no formal schooling to a bachelor’s degree. A total of 75.3% of the children with clubfoot were boys, resided in a city, and were older than five months. In terms of medical conditions, 60% of the children had clubfoot on both feet, while 70.6% of the neonates were born by cesarean section. 76.5% of the people were in the bracing phase of Ponseti treatment, and 94.1% of them had no difficulties with their treatment. 21.2% of caregivers had high blood pressure, while 57.6% did not have any other health problems. Only 10.6% of the children had other health problems as well. Using the DASS-21 scale, a psychological test found that 50.6% of caregivers stated they were worried (48.8% extremely severe), 28.2% said they were stressed (45.8% seriously stressed), and 44.7% said they were depressed (26.3% extremely severe).

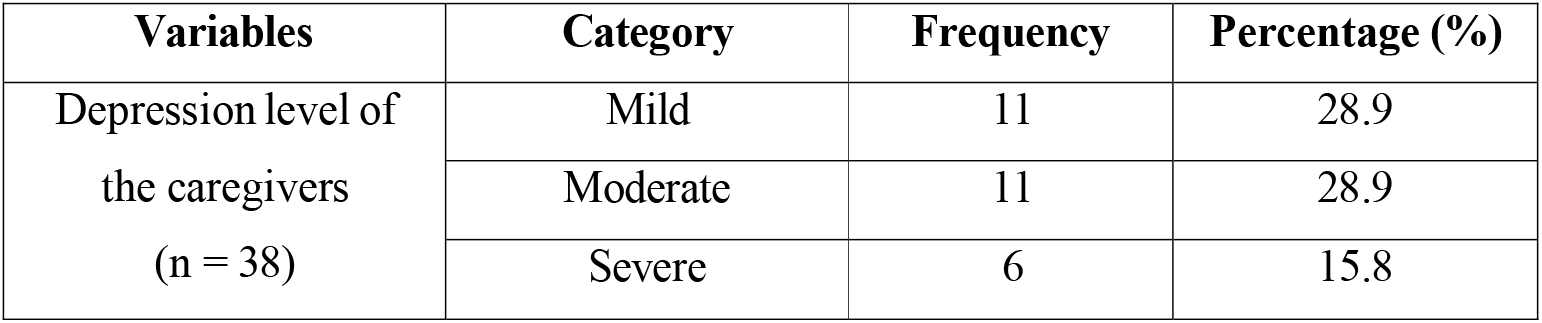
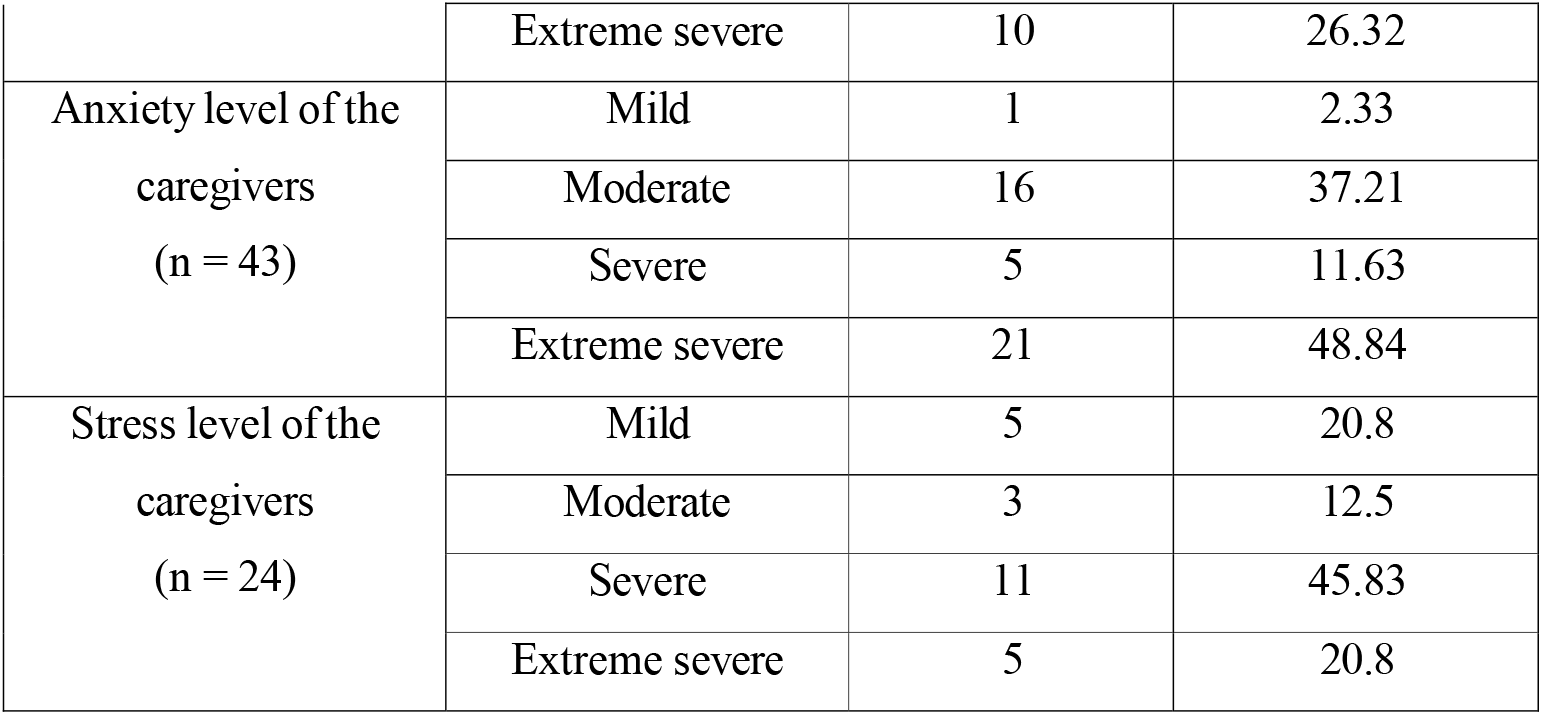

Statistical analysis showed that monthly income was obviously connected to anxiety (p=0.009), whereas education level was strongly linked to depression (p=0.029). The child’s birth weight (p=0.013), the caregiver’s age (p=0.053), and the way the baby was delivered (p=0.056) all had some effect on stress.

| Dependent Variable | Significant Independent Variable | Value | p-Value |
| --- | --- | --- | --- |
| **Depression** | Education of caregiver | 22.807 | 0.029** |
| **Anxiety** | Monthly income | 26.594 | 0.009** |
| **Stress** | Age group of caregiver | 20.818 | 0.053* |
|  | Birth weight | 31.239 | 0.013** |
|  | Mode of delivery | 9.231 | 0.056* |

\*→ Marginally significant (p < 0.10), \*\* → Statistically significant (p < 0.05), \*\*\* → Highly significant (p < 0.01)

There was no clear link between psychological discomfort and other criteria like gender, job, or where the person lived. These results demonstrate that taking care of a child with clubfoot can be very bad for mental health, especially for people who are less educated, have less money, or are caring for babies who were born too small or after a cesarean section. This shows that caretakers of people with clubfoot require more than just medical care; they also need emotional assistance. Statistical tests showed that monthly income was obviously associated to anxiety (p=0.009), whereas education level was strongly linked to depression (p=0.029). The child’s birth weight (p=0.013), the caregiver’s age (p=0.053), and the way the baby was delivered (p=0.056) all had some effect on stress.

## Discussion

This study backs up what other research has found: parents of kids getting Ponseti treatment for congenital clubfoot are quite unhappy, with feelings of melancholy, anxiety, and tension. Dong et al. say that mothers’ anxiety and sadness are higher at the beginning of treatment and usually go back to normal after therapy [11, 8]. In the same way, Mohandas et al. talked about the mental health problems that parents of kids with genetic abnormalities face and how these affect the whole family [12, 16]. During the bracing period, caregivers’ quality of life also suffers. Besselaar et al. say that kids with braces have physical pain and trouble sleeping, which may make caregivers more stressed [12, 3]. Parents’ worry grew since their kids were complaining about being irritable and not sleeping well while wearing braces.

Child resistance and logistical problems often made it hard to follow the orthosis, which is in line with what Sassi et al. found, which showed that caregivers have a hard time keeping up with brace use [13, 16]. Drew’s ethnographic study looked at systemic problems that make it harder for people to stick to their treatment and put more stress on caregivers, such as not having enough resources and not being able to get to the clinic easily [14,7]. Help with schoolwork was connected to less stress and more confidence in caregivers. Parents who were given clear information on the Ponseti approach showed better understanding and adaptability, which is in line with Ali’s research on the benefits of educational materials [15,16] and Rasheed et al.’s focus on parental education [17]. The pain that caregivers felt during casting made their worry even worse. Upadhyay et al. showed that non-drug pain management methods, such giving neonates sucrose, can help them feel better while they are being cast [18, 6, 9]. Using these methods might make things easier for kids and their caregivers.

In the end, mistakes made during therapy make caretakers nervous and hurt results. Youn et al. stressed the need for strict staff training and following guidelines in the Ponseti strategy to keep caregivers’ trust and get the best results [20]. There is a strong link between how effectively treatment works and how mentally well caretakers are. To improve adherence and psychosocial outcomes, we need to deal with logistical problems, provide people information, help them deal with pain, and make sure they stick to their treatment. Using these strategies together can improve family-centered care for treating clubfoot.

## Conclusion

The study looked at the mental impacts of taking care of CTEV children and found that low income, low birth weight, and the way the child was born all made people more stressed, anxious, and sad. Even while CTEV didn’t have much of an effect on social or family life, money problems, notably the cost of traveling for rural families,-guys, were big. The results show that caregivers require decentralized services to make their lives easier and better. Studies that looked at both sociodemographic characteristics and psychological distress found a correlation between the two. In future studies, random sampling, bigger sample numbers, and longer study periods will make the results more generalizable. It is important to use recognized methods to measure psychological discomfort, cognitive problems, and sleep problems. To completely understand the problems caregivers face and come up with CTEV therapy and assistance, future research should include caregivers from all around Bangladesh.

## Data Availability

All data produced in the present study are available upon reasonable request to the authors

